# IDENTIFYING DIFFERENT PHENOTYPES OF SYMPTOMATIC GASTROESOPHAGEAL REFLUX DISEASE USING ARTIFICIAL INTELLIGENCE

**DOI:** 10.1101/2023.04.14.23288596

**Authors:** Jerry D. Gardner, George Triadafilopoulos

## Abstract

**Introduction:** The Lyon Consensus Conference proposed criteria for the clinical diagnosis of three different phenotypes of gastroesophageal reflux disease: nonerosive gastroesophageal reflux disease, Reflux Hypersensitivity, and Functional Heartburn.

**Methods:** In the present study, we examined the potential of ChatGPT, an artificial intelligence-based conversational large language model to describe how one can identify the different phenotypes as identified by the Lyon Consensus Conference, and to provide a diagnosis when given important clinical findings in a given patient with a particular phenotype.

**Results:** Although in our analyses ChatGPT captured correct information regarding symptoms, upper gastrointestinal endoscopy findings and response to gastric antisecretory agents when asked to describe different phenotypes, it failed, however, to return correct information on esophageal acid exposure time and the association of symptoms with esophageal reflux episodes. ChatGPT was even less effective in returning the correct diagnosis after being given specific clinical features of a particular phenotype.

**Conclusions:** Although it seems likely that the ability of ChatGPT to capture information from multiple sources will improve with future use and refinement, presently it is inadequate as a standalone tool for processing information for the description or diagnosis of different clinical disease states. On the other hand, artificial intelligence might prove useful to clinicians in performing tasks that involve obtaining data from a single source such as the electronic medical record and generating a document having a standardized format such as a discharge summary.

## INTRODUCTION

The Lyon Consensus Conference (1, 2) proposed criteria for the clinical diagnosis of three different phenotypes of gastroesophageal reflux disease (GERD): nonerosive gastroesophageal reflux disease (NERD), Reflux Hypersensitivity, and Functional Heartburn.

Functional Heartburn subjects and Reflux Hypersensitivity subjects have normal esophageal acid exposure time (AET) with esophageal pH <4 for less than 4% of the 24-hour esophageal pH recording. NERD subjects have increased esophageal AET of pH <4 for greater than 6% of the 24-hour esophageal pH recording. Reflux Hypersensitivity subjects have a positive association of symptoms with reflux episodes (symptom index SI (3) and symptom-association probability SAP (4)), whereas Functional Heartburn subjects have no association of symptoms with reflux episodes (negative SI and negative SAP).

## METHODS

In the present study, we examined the potential of ChatGPT (5, 6), an artificial intelligence (AI)-based conversational large language model to describe how one can identify the different GERD phenotypes as defined identified by the Lyon Consensus Conference, and to provide a diagnosis when given important clinical findings in a given patient with a particular GERD phenotype.

We chose GERD phenotypes to explore the potential of ChatGPT because the phenotypes share esophageal reflux symptoms and a normal upper gastrointestinal endoscopy, but differ in AET and the association of symptoms with esophageal reflux episodes (1, 2). We were particularly interested in whether the AI could report the similarities as well as the differences in the phenotypes.

## RESULTS

The verbatim questions and responses from ChatGPT are given in APPENDIX.

Table 1 summarizes the responses given by ChatGPT to questions regarding GERD or one of the three GERD phenotypes. ChatGPT consistently provided correct information regarding symptoms and findings on upper gastrointestinal endoscopy, but failed to provide information regarding esophageal acid exposure time and association of symptoms with esophageal reflux events (SAP and SI). ChatGPT provided correct information regarding possible response to proton pump inhibitors or histamine H2 receptor antagonists for GERD, NERD and Reflux Hypersensitivity, but did not respond that Functional Heartburn typically does not respond to gastric antisecretory agents.

**TABLE 1.** FINDINGS MENTIONED BY ChatGPT IN RESPONSE TO QUESTION.

| **CLINICAL FINDINGS** | **YES/NO** |
|---|---|
| WHAT IS GERD? |  |
| SYMPTOMS – HEARTBURN, REGURGITATION, CHEST PAIN | YES |
| UPPER GI ENDOSCOPY MAY SHOW ESOPHAGITIS | YES |
| AET >6.0 | NO |
| SAP AND SI POSITIVE OR NEGATIVE | NO |
| SYMPTOMS AND ESOPHAGITIS RESPOND TO PPIs OR H2RAs | YES |
| WHAT IS NERD? |  |
| SYMPTOMS – HEARTBURN, REGURGITATION, CHEST PAIN | YES |
| NORMAL UPPER GI ENDOSCOPY | YES |
| AET >6.0 | NO |
| SAP AND SI POSITIVE OR NEGATIVE | NO |
| SYMPTOMS RESPOND TO PPIs OR H2RAs | YES |
| WHAT IS REFLUX HYPERSENSITIVITY? |  |
| SYMPTOMS – HEARTBURN, REGURGITATION, CHEST PAIN | YES |
| NORMAL UPPER GI ENDOSCOPY | YES |
| AET< 4.0 | NO |
| SAP AND SI POSITIVE | NO |
| SYMPTOMS SOMETIMES RESPOND TO PPIs OR H2RAs | YES |
| WHAT IS FUNCTIONAL HEARTBURN? |  |
| SYMPTOMS – HEARTBURN, REGURGITATION, CHEST PAIN | YES |
| NORMAL UPPER GI ENDOSCOPY | YES |
| AET < 4.0 | NO |
| SAP AND SI NEGATIVE | NO |
| SYMPTOMS USUALLY DO NOT RESPOND TO PPIs OR H2RAs | NO |

The following section gives the responses of ChatGPT when asked to provide a diagnosis after being presented with a patient’s clinical findings.

## DIAGNOSES

### GERD

#### WHAT IS DIAGNOSIS?

50 YO MALE. HEARTBURN, REGURGITATION, CHEST PAIN FOR 6 MONTHS. UPPER GI ENDOSCOPY SHOWS ESOPHAGITIS. AET 8.5 %. SAP AND SI POSITIVE. SYMPTOMS AND ESOPHAGITIS RESPOND TO PPI.

### ChatGPT -- GERD

#### WHAT IS DIAGNOSIS?

50 YO MALE. HEARTBURN, REGURGITATION, CHEST PAIN FOR 6 MONTHS. UPPER GI ENDOSCOPY SHOWS ESOPHAGITIS. AET 8.5 %. SAP AND SI NEGATIVE. SYMPTOMS AND ESOPHAGITIS RESPOND TO PPI.

### ChatGPT -- NERD

#### WHAT IS DIAGNOSIS?

50 YO MALE. HEARTBURN, REGURGITATION, CHEST PAIN FOR 6 MONTHS. UPPER GI ENDOSCOPY SHOWS ESOPHAGITIS. AET 8.5 %. SAP AND SI POSITIVE. SYMPTOMS AND ESOPHAGITIS SO NOT RESPOND TO PPI.

### ChatGPT -- GERD

#### WHAT IS DIAGNOSIS?

50 YO MALE. HEARTBURN, REGURGITATION, CHEST PAIN FOR 6 MONTHS. UPPER GI ENDOSCOPY SHOWS ESOPHAGITIS. AET 8.5 %. SAP AND SI NEGATIVE. SYMPTOMS AND ESOPHAGITIS DO NOT RESPOND TO PPI.

### ChatGPT – UNKNOWN

When asked the diagnosis after being given clinical characteristics of four typical GERD patients, ChatGPT correctly identified two patients, misidentified one patient as NERD, and was unable to identify one patient.

### NERD

#### WHAT IS DIAGNOSIS?

50 YO MALE. HEARTBURN, REGURGITATION, CHEST PAIN FOR 6 MONTHS. UPPER GI ENDOSCOPY IS NORMAL. AET 8.5 %. SAP AND SI POSITIVE. SYMPTOMS RESPOND TO PPI.

### ChatGPT -- GERD. MISINTERPRETED SI AND SAP AS SCHILLING TEST AND SODIUM ACID PHOSPHATE, RESPECTIVELY

#### WHAT IS DIAGNOSIS?

50 YO MALE. HEARTBURN, REGURGITATION, CHEST PAIN FOR 6 MONTHS. UPPER GI ENDOSCOPY IS NORMAL AET 8.5 %. SAP AND SI NEGATIVE. SYMPTOMS RESPOND TO PPI.

### ChatGPT -- NOT DUE TO REFLUX. MISINTERPRETED SI AND SAP AS SCHILLING TEST AND SODIUM ACID PHOSPHATE, RESPECTIVELY

#### WHAT IS DIAGNOSIS?

50 YO MALE. HEARTBURN, REGURGITATION, CHEST PAIN FOR 6 MONTHS. UPPER GI ENDOSCOPY IS NORMAL AET 8.5 %. SAP AND SI POSITIVE. SYMPTOMS DO NOT RESPOND TO PPI.

### ChatGPT -- NOT DUE TO GERD. MISINTERPRETED SI AND SAP AS SCHILLING TEST AND SODIUM ACID PHOSPHATE, RESPECTIVELY

#### WHAT IS DIAGNOSIS?

50 YO MALE. HEARTBURN, REGURGITATION, CHEST PAIN FOR 6 MONTHS. UPPER GI ENDOSCOPY IS NORMAL. AET 8.5 %. SAP AND SI NEGATIVE. SYMPTOMS DO NOT RESPOND TO PPI.

### ChatGPT -- NOT DUE TO GERD. MISINTERPRETED SI AND SAP AS SCHILLING TEST AND SODIUM ACID PHOSPHATE, RESPECTIVELY

When asked the diagnosis after being given clinical characteristics of four typical NERD patients, ChatGPT identified one patient as GERD, and was unable to identify the other three patients. ChatGPT also misidentified the SA and SAP as Schilling test and sodium acid phosphate in all four patients.

### REFLUX HYPERSENSITIVITY

#### WHAT IS DIAGNOSIS?

50 YO MALE. HEARTBURN, REGURGITATION, CHEST PAIN FOR 6 MONTHS. UPPER GI ENDOSCOPY IS NORMAL. AET 3.3 %. SAP AND SI POSITIVE. SYMPTOMS RESPOND TO PPI.

### ChatGPT -- GERD WITH HYPERSENSITIVITY COMPONENT

#### WHAT IS DIAGNOSIS?

50 YO MALE. HEARTBURN, REGURGITATION, CHEST PAIN FOR 6 MONTHS. UPPER GI ENDOSCOPY IS NORMAL AET 3.3 %. SAP AND SI POSITIVE. SYMPTOMS DO NOT RESPOND TO PPI.

#### ChatGPT -- NON-ACID REFLUX OR ANOTHER CONDITION

When asked the diagnosis after being given clinical characteristics of two typical patients with Reflux Hypersensitivity, ChatGPT correctly identified one patient, but was unable to make a diagnosis for the other patient.

### FUNCTIONAL HEARTBURN

#### WHAT IS DIAGNOSIS?

50 YO MALE. HEARTBURN, REGURGITATION, CHEST PAIN FOR 6 MONTHS. UPPER GI ENDOSCOPY IS NORMAL. AET 3.3 %. SAP AND SI NEGATIVE. SYMPTOMS RESPOND TO PPI.

### ChatGPT -- GERD

#### WHAT IS DIAGNOSIS?

50 YO MALE. HEARTBURN, REGURGITATION, CHEST PAIN FOR 6 MONTHS. UPPER GI ENDOSCOPY IS NORMAL AET 3.3 %. SAP AND SI NEGATIVE. SYMPTOMS DO NOT RESPOND TO PPI.

### ChatGPT -- NOT DUE TO GERD. MISINTERPRETED SI AND SAP AS SCHILLING TEST AND SODIUM ACID PHOSPHATE, RESPECTIVELY

When asked the diagnosis after being given clinical characteristics of two typical patients with Functional Heartburn, ChatGPT misidentified one patient as GERD, and was unable to identify the other patient. As above ChatGPT also misidentified the SA and SAP as Schilling test and sodium acid phosphate in one patient.

## DISCUSSION

In making decisions regarding diagnosis and treatment, clinicians routinely access multiple sources of data. The present analyses illustrate that in describing a GERD phenotype, ChatGPT must consider a particular patient’s symptoms, the results from an upper gastrointestinal endoscopy, the esophageal AET calculated from an esophageal pH recording, the SI and SAP, both of which are calculated from the time of a symptom and the associated gastroesophageal reflux episode observed with esophageal impedance testing or measurement of esophageal pH, and the response of symptoms and abnormalities on endoscopy to gastric antisecretory agents. A review of the abilities of various AI models to capture data from multiple sources described this area of research as being “relatively underdeveloped” (7, 8).

Although in our analyses ChatGPT captured correct information regarding symptoms, upper gastrointestinal endoscopy findings and response to gastric antisecretory agents when asked to describe different GERD phenotypes, it failed to return correct information on esophageal AET and the association of symptoms with esophageal reflux episodes. ChatGPT was even less effective in returning the correct diagnosis after being given specific clinical features. The AI returned two out of four correct responses for a GERD diagnosis, and only one out of eight correct responses for GERD phenotypes. Although it seems likely that the ability of ChatGPT to capture information from multiple sources will improve with future use and refinement, presently it is inadequate as a standalone tool for processing information for the description or diagnosis of different clinical disease states.

On the other hand, AI might prove useful to clinicians in performing tasks that involve obtaining data from a single source such as the electronic medical record and generating a document having a standardized format such as a discharge summary. AI might also be useful to researchers who perform meta-analyses by extracting similar data from many publications and performing standardized presentations and analyses of the data.

One possible, albeit unlikely explanation for the poor performance of ChatGPT in describing or diagnosing the various GERD phenotypes is that it is due to limitations in the Lyon Consensus process. For example, the failure of ChatGPT to return correct information regarding esophageal acid exposure time and the association of symptoms with esophageal reflux episodes in describing GERD phenotypes may be due insufficient data in the medical literature to include these measures in the description. Similarly, the poor performance of ChatGPT in diagnosing various GERD phenotypes might reflect limitations in available published data supporting the diagnosis of a particular GERD phenotype. Although all consensus processes have limitations, it seems highly unlikely that the performance of ChatGPT reflects important limitations in the Lyon Consensus process because it involved an international panel of GERD experts who reviewed the published literature and graded the evidence regarding GERD phenotypes over a period of at least two years before generating the consensus statements (1).

AI systems that learn patterns directly from raw data have made substantial progress in interpreting various types of medical images and are being incorporated into the daily practice of many medical specialties, including gastroenterology, that routinely rely on the interpretation of images (8). In other instances, AI systems require labels and inputs of data into appropriate labels to train the system (8, 9). This process can be particularly labor-intensive and is subject to misinterpretation as evidenced by ChatGPT misidentifying the SI as Schilling test and SAP as sodium acid phosphate.

Although many examinations of AI systems have compared the performance of a particular system to that of a human (8, 10), it seems likely that an important future use of AI will be in partnership with a medical clinician or researcher. For example, professional organizations routinely generate guidelines for practitioners and incorporate AI algorithms into the development process. As such, they might identify areas where the evidence is of poor quality or point out important information that needs to be considered.

## Data Availability

All data produced in the present work are contained in the manuscript

## APPENDIX

### VERBATIM RESPONSES FROM ChatGPT

#### What is GERD?

GERD stands for Gastroesophageal Reflux Disease, a chronic digestive condition in which the stomach’s contents, including acid, flow back into the esophagus. This can cause a range of symptoms, including heartburn, regurgitation, and difficulty swallowing, among others. GERD can be caused by a number of factors, including obesity, pregnancy, certain foods and drinks, and certain medications. In severe cases, GERD can lead to more serious health problems, such as esophagitis, Barrett’s esophagus, and even esophageal cancer. Treatment options for GERD typically involve lifestyle changes, such as losing weight, avoiding trigger foods and drinks, and avoiding lying down immediately after eating, as well as medications, such as antacids, H2 receptor blockers, and proton pump inhibitors. In some cases, surgery may be recommended to help manage the symptoms of GERD.

#### What is NERD?

NERD stands for Non-erosive Reflux Disease. It is a subtype of Gastroesophageal Reflux Disease (GERD), which is a chronic digestive condition in which the contents of the stomach, including acid, flow back into the esophagus. NERD is characterized by the presence of symptoms typical of GERD, such as heartburn and regurgitation, but without any evidence of damage to the lining of the esophagus. This is in contrast to erosive GERD, in which there is damage to the esophagus. The exact cause of NERD is not well understood, but it is thought to be related to factors such as hiatal hernias, weakened lower esophageal sphincter function, and certain lifestyle factors, such as obesity and smoking. Treatment for NERD typically involves lifestyle changes, such as losing weight, avoiding trigger foods and drinks, and avoiding lying down immediately after eating, as well as medications, such as antacids, H2 receptor blockers, and proton pump inhibitors. In some cases, surgery may be recommended to help manage the symptoms of GERD.

#### What is reflux hypersensitivity?

Reflux hypersensitivity is a condition in which individuals experience symptoms of acid reflux, such as heartburn and regurgitation, in response to normal levels of gastroesophageal reflux. This means that even small amounts of acid reflux can cause symptoms, even though there is no evidence of tissue damage. It is thought that reflux hypersensitivity may be due to an abnormal sensitivity of the esophagus to the presence of acid. It can also be related to other factors, such as stress, anxiety, and depression. Reflux hypersensitivity is distinct from Gastroesophageal Reflux Disease (GERD), in which there is damage to the esophagus from repeated exposure to acid reflux. The treatment for reflux hypersensitivity may include lifestyle changes, such as avoiding trigger foods and drinks, avoiding lying down immediately after eating, and managing stress, as well as medications, such as antacids, H2 receptor blockers, and proton pump inhibitors. In some cases, referral to a psychologist or psychiatrist may be recommended to help manage symptoms related to stress, anxiety, and depression.

#### What is functional heartburn?

Functional heartburn is a type of indigestion that is characterized by symptoms of heartburn and acid reflux, but without any identifiable structural or biochemical abnormality in the gastrointestinal tract. In other words, functional heartburn is a condition in which the symptoms of heartburn and acid reflux are not due to any underlying physical problem with the esophagus or stomach.

Functional heartburn is often related to lifestyle factors, such as stress, anxiety, and depression, as well as other factors, such as certain foods and drinks, and medications. It is also thought to be related to abnormalities in the function of the lower esophageal sphincter, which is the muscle that prevents acid from flowing back into the esophagus.

Treatment for functional heartburn may involve lifestyle changes, such as avoiding trigger foods and drinks, avoiding lying down immediately after eating, and managing stress, as well as medications, such as antacids, H2 receptor blockers, and proton pump inhibitors. In some cases, referral to a psychologist or psychiatrist may be recommended to help manage symptoms related to stress, anxiety, and depression.

#### WHAT IS DIAGNOSIS? 50 YO MALE. HEARTBURN, REGURGITATION, CHEST PAIN FOR 6 MONTHS. UPPER GI ENDOSCOPY SHOWS ESOPHAGITIS. AET 8.5 %. SAP AND SI POSITIVE. SYMPTOMS AND ESOPHAGITIS RESPOND TO PPI

The diagnosis in this case is likely to be Gastroesophageal Reflux Disease (GERD). This is based on the symptoms of heartburn, regurgitation, and chest pain that have been present for 6 months and have been confirmed by an upper GI endoscopy which showed esophagitis. The esophageal acid exposure time (AET) of 8.5% and the positive results for acid and non-acid reflux on the concurrent 24-hour esophageal pH-impedance monitoring (SAP and SI) also support the diagnosis of GERD. Additionally, the fact that the symptoms and esophagitis have responded to proton pump inhibitor (PPI) treatment also supports the diagnosis of GERD.

#### WHAT IS DIAGNOSIS? 50 YO MALE. HEARTBURN, REGURGITATION, CHEST PAIN FOR 6 MONTHS. UPPER GI ENDOSCOPY SHOWS ESOPHAGITIS. AET 8.5 %. SAP AND SI NEGATIVE. SYMPTOMS AND ESOPHAGITIS RESPOND TO PPI

The diagnosis in this case is likely to be non-erosive reflux disease (NERD). This is based on the symptoms of heartburn, regurgitation, and chest pain that have been present for 6 months and have been confirmed by an upper GI endoscopy which showed esophagitis. However, the fact that the esophageal acid exposure time (AET) was only 8.5% and that the results for acid and non-acid reflux on the concurrent 24-hour esophageal pH-impedance monitoring (SAP and SI) were negative suggest that the esophagitis is not caused by acid reflux. Additionally, the fact that the symptoms and esophagitis have responded to proton pump inhibitor (PPI) treatment also supports the diagnosis of NERD.

#### WHAT IS DIAGNOSIS? 50 YO MALE. HEARTBURN, REGURGITATION, CHEST PAIN FOR 6 MONTHS. UPPER GI ENDOSCOPY SHOWS ESOPHAGITIS. AET 8.5 %. SAP AND SI POSITIVE. SYMPTOMS AND ESOPHAGITIS SO NOT RESPOND TO PPI

The diagnosis in this case may be complicated gastroesophageal reflux disease (GERD). The symptoms of heartburn, regurgitation, and chest pain for 6 months, confirmed by an upper GI endoscopy showing esophagitis, along with a positive result for acid and non-acid reflux on concurrent 24-hour esophageal pH-impedance monitoring (SAP and SI) and an esophageal acid exposure time (AET) of 8.5% are all consistent with GERD. However, the fact that the symptoms and esophagitis did not respond to proton pump inhibitor (PPI) treatment suggests that there may be additional factors contributing to

#### WHAT IS DIAGNOSIS? 50 YO MALE. HEARTBURN, REGURGITATION, CHEST PAIN FOR 6 MONTHS. UPPER GI ENDOSCOPY SHOWS ESOPHAGITIS. AET 8.5 %. SAP AND SI NEGATIVE. SYMPTOMS AND ESOPHAGITIS DO NOT RESPOND TO PPI

The diagnosis in this case may be difficult to determine based on the available information. The symptoms of heartburn, regurgitation, and chest pain for 6 months, confirmed by an upper GI endoscopy showing esophagitis, and a low esophageal acid exposure time (AET) of 8.5% with negative results for acid and non-acid reflux on concurrent 24-hour esophageal pH-impedance monitoring (SAP and SI) suggest that acid reflux may not be the primary cause of the symptoms. However, the fact that the symptoms and esophagitis did not respond to proton pump inhibitor (PPI) treatment makes it difficult to determine the underlying cause. Further evaluation and testing may be necessary to determine the cause of the symptoms and the most appropriate course of treatment.

This could include additional endoscopic evaluations, pH monitoring, or even biopsy of the esophagus.

#### NERD WHAT IS DIAGNOSIS? 50 YO MALE. HEARTBURN, REGURGITATION, CHEST PAIN FOR 6 MONTHS. UPPER GI ENDOSCOPY IS NORMAL. AET 8.5 %. SAP AND SI POSITIVE. SYMPTOMS RESPOND TO PPI

The patient is a 50-year-old male with symptoms of heartburn, regurgitation, and chest pain for the past 6 months. An upper gastrointestinal endoscopy was performed and was found to be normal. An AET (Acid Exposure Time) test was performed and showed a result of 8.5%. A SAP (Sodium Acid Phosphate) test and an SI (Schilling Test) were both positive. The patient’s symptoms respond to proton pump inhibitors (PPIs).

Based on this information, it appears that the patient may have gastroesophageal reflux disease (GERD). GERD is a condition where stomach acid backs up into the esophagus, causing symptoms such as heartburn and regurgitation. The AET and SAP results suggest that there is an increase in acid exposure in the esophagus, which is consistent with GERD. The positive response to PPIs further supports this diagnosis. However, a final diagnosis should only be made by a healthcare professional after further evaluation and investigation.

#### WHAT IS DIAGNOSIS? 50 YO MALE. HEARTBURN, REGURGITATION, CHEST PAIN FOR 6 MONTHS. UPPER GI ENDOSCOPY IS NORMAL AET 8.5 %. SAP AND SI NEGATIVE. SYMPTOMS RESPOND TO PPI

The patient is a 50-year-old male with symptoms of heartburn, regurgitation, and chest pain for the past 6 months. An upper gastrointestinal endoscopy was performed and was found to be normal. An AET (Acid Exposure Time) test was performed and showed a result of 8.5%. A SAP (Sodium Acid Phosphate) test and an SI (Schilling Test) were both negative. The patient’s symptoms respond to proton pump inhibitors (PPIs).

Based on this information, the diagnosis is less clear. The normal upper GI endoscopy and negative SAP and SI results suggest that there may not be an issue with acid reflux. However, the positive response to PPIs and the presence of symptoms such as heartburn and regurgitation could still indicate gastroesophageal reflux disease (GERD). Additionally, chest pain can be a symptom of other conditions such as heart disease, so a thorough evaluation is necessary to determine the cause of the patient’s symptoms. A final diagnosis should only be made by a healthcare professional after further investigation and testing.

#### WHAT IS DIAGNOSIS? 50 YO MALE. HEARTBURN, REGURGITATION, CHEST PAIN FOR 6 MONTHS. UPPER GI ENDOSCOPY IS NORMAL AET 8.5 %. SAP AND SI POSITIVE. SYMPTOMS DO NOT RESPOND TO PPI

The patient is a 50-year-old male with symptoms of heartburn, regurgitation, and chest pain for the past 6 months. An upper gastrointestinal endoscopy was performed and was found to be normal. An AET (Acid Exposure Time) test was performed and showed a result of 8.5%. A SAP (Sodium Acid Phosphate) test and an SI (Schilling Test) were both positive. The patient’s symptoms do not respond to proton pump inhibitors (PPIs).

This information suggests that the patient may have a condition other than gastroesophageal reflux disease (GERD). GERD is typically treated with PPIs and a positive response to these medications is usually seen in patients with GERD. However, in this case, the patient’s symptoms do not respond to PPIs, which raises the possibility of another underlying condition. Chest pain can also be a symptom of conditions such as heart disease, so a thorough evaluation is necessary to determine the cause of the patient’s symptoms. A final diagnosis should only be made by a healthcare professional after further investigation and testing.

#### WHAT IS DIAGNOSIS? 50 YO MALE. HEARTBURN, REGURGITATION, CHEST PAIN FOR 6 MONTHS. UPPER GI ENDOSCOPY IS NORMAL. AET 8.5 %. SAP AND SI NEGATIVE. SYMPTOMS DO NOT RESPOND TO PPI

The patient is a 50-year-old male with symptoms of heartburn, regurgitation, and chest pain for the past 6 months. An upper gastrointestinal endoscopy was performed and was found to be normal. An

AET (Acid Exposure Time) test was performed and showed a result of 8.5%. A SAP (Sodium Acid Phosphate) test and an SI (Schilling Test) were both negative. The patient’s symptoms do not respond to proton pump inhibitors (PPIs).

This information makes it difficult to determine a clear diagnosis. The normal upper GI endoscopy and negative SAP and SI results suggest that there may not be an issue with acid reflux, which is typically treated with PPIs. However, the patient’s symptoms of heartburn and regurgitation, and the fact that the symptoms do not respond to PPIs, could indicate that there is another underlying condition causing the symptoms. Chest pain can also be a symptom of conditions such as heart disease, so a thorough evaluation is necessary to determine the cause of the patient’s symptoms. A final diagnosis should only be made by a healthcare professional after further investigation and testing.

#### REFLUX HYPERSENSITIVITY WHAT IS DIAGNOSIS? 50 YO MALE. HEARTBURN, REGURGITATION, CHEST PAIN FOR 6 MONTHS. UPPER GI ENDOSCOPY IS NORMAL. AET 3.3 %. SAP AND SI POSITIVE. SYMPTOMS RESPOND TO PPI

Based on the symptoms and test results described, it appears that the individual may have been diagnosed with gastroesophageal reflux disease (GERD) with a hypersensitivity component.

The individual is a 50-year-old male who has been experiencing heartburn, regurgitation, and chest pain for 6 months. An upper GI endoscopy was performed and was found to be normal. The AET (Acid Exposure Time) was found to be 3.3%. The SAP (symptom association probability) and SI (symptom index) were both positive, indicating a strong correlation between the symptoms and acid reflux. The individual’s symptoms have responded well to proton pump inhibitors (PPIs), which are commonly used to treat GERD.

In conclusion, the diagnosis of GERD with a hypersensitivity component is likely based on the individual’s symptoms, test results, and response to treatment with PPIs.

#### WHAT IS DIAGNOSIS? 50 YO MALE. HEARTBURN, REGURGITATION, CHEST PAIN FOR 6 MONTHS. UPPER GI ENDOSCOPY IS NORMAL AET 3.3 %. SAP AND SI POSITIVE. SYMPTOMS DO NOT RESPOND TO PPI

Based on the updated information, it appears that the individual’s symptoms may not be solely due to gastroesophageal reflux disease (GERD). The individual is a 50-year-old male who has been experiencing heartburn, regurgitation, and chest pain for 6 months. An upper GI endoscopy was performed and was found to be normal. The AET (Acid Exposure Time) was found to be 3.3%. The SAP (symptom association probability) and SI (symptom index) were both positive, indicating a strong correlation between the symptoms and acid reflux. However, the individual’s symptoms did not respond to proton pump inhibitors (PPIs), which are commonly used to treat GERD.

Given the normal results of the upper GI endoscopy and the lack of response to PPIs, it is possible that the individual may have non-acid reflux or another condition that is causing their symptoms.

Further evaluation, such as additional tests or referral to a specialist, may be necessary to determine the underlying cause of the individual’s symptoms and make a more accurate diagnosis.

#### WHAT IS DIAGNOSIS? 50 YO MALE. HEARTBURN, REGURGITATION, CHEST PAIN FOR 6 MONTHS. UPPER GI ENDOSCOPY IS NORMAL. AET 3.3 %. SAP AND SI NEGATIVE. SYMPTOMS RESPOND TO PPI

Based on the information provided, the diagnosis for this 50-year-old male who has experienced heartburn, regurgitation, and chest pain for 6 months and has normal results from an upper GI endoscopy with a normal amount of acid exposure and negative results from a test for swallowing problems, the symptoms suggest gastroesophageal reflux disease (GERD). The positive response to proton pump inhibitor (PPI) treatment further supports this diagnosis. However, it’s important to note that this information is limited, and a full and thorough evaluation by a medical professional is necessary to make a definitive diagnosis.

#### WHAT IS DIAGNOSIS? 50 YO MALE. HEARTBURN, REGURGITATION, CHEST PAIN FOR 6 MONTHS. UPPER GI ENDOSCOPY IS NORMAL AET 3.3 %. SAP AND SI NEGATIVE. SYMPTOMS DO NOT RESPOND TO PPI

In this case, the fact that the patient’s symptoms of heartburn, regurgitation, and chest pain have persisted for 6 months despite treatment with proton pump inhibitors (PPIs) suggests that a different diagnosis may need to be considered. The normal results of the upper GI endoscopy and the normal acid exposure time (AET) of 3.3% suggest that gastroesophageal reflux disease (GERD) may not be the underlying cause of the patient’s symptoms. The negative results of the swallowing test (SAP and SI) also support this.

Given that the patient’s symptoms do not respond to PPIs, further testing and evaluation may be necessary to determine the cause of the symptoms and make an accurate diagnosis. This may include additional tests such as barium swallow, pH monitoring, or further imaging studies. It’s important for the patient to consult with their healthcare provider for a complete evaluation and to develop an appropriate treatment plan.

## Notes

**Conflict of Interest Statement:** Neither author has a conflict of interest with the analyses in the present paper.

### Competing Interest Statement

The authors have declared no competing interest.

### Funding Statement

This study did not receive any funding

